# Comprehensive reanalysis for CNVs in ES data from unsolved rare disease cases results in new diagnoses

**DOI:** 10.1101/2023.10.22.23296993

**Authors:** German Demidov, Burcu Yaldiz, José Garcia-Pelaez, Elke de Boer, Nika Schuermans, Liedewei Van de Vondel, Ida Paramonov, Lennart F. Johansson, Francesco Musacchia, Elisa Benetti, Gemma Bullich, Karolis Sablauskas, Sergi Beltran, Christian Gilissen, Alexander Hoischen, Stephan Ossowski, Richarda de Voer, Katja Lohmann, Carla Oliveira, Ana Topf, Lisenka E.L.M. Vissers, the Solve-RD Consortia, Steven Laurie

## Abstract

We report the diagnostic results of a comprehensive copy number variant (CNV) reanalysis of 9,171 exome sequencing (ES) datasets from 5,757 families, including 6,143 individuals affected by a rare disease (RD). The data analysed was extremely heterogeneous, having been generated using 28 different exome enrichment kits, and sequenced on multiple short-read sequencing platforms, by 42 different research groups across Europe partnering in the Solve-RD project. Each of these research groups had previously undertaken their own analysis of the ES data but had failed to identify disease-causing variants.

We applied three CNV calling algorithms to maximise sensitivity: ClinCNV, Conifer, and ExomeDepth. Rare CNVs overlapping genes of interest in custom lists provided by one of four partner European Reference Networks (ERN) were identified and taken forward for interpretation by clinical experts in RD. To facilitate interpretation, Integrative Genomics Viewer (IGV) screenshots incorporating a variety of custom-made tracks were generated for all prioritised CNVs.

These analyses have resulted in a molecular diagnosis being provided for 51 families in this sample, with ClinCNV performing the best of the three algorithms in identifying disease-causing CNVs. We also identified pathogenic CNVs that are partially explanatory of the proband’s phenotype in a further 34 individuals. This work illustrates the value of reanalysing ES *cold cases* for CNVs even where analyses had been undertaken previously. Crucially, identification of these previously undetected CNVs has resulted in the conclusion of the diagnostic odyssey for these RD families, some of which had endured decades.

## Introduction

Rare diseases (RD) are defined in Europe as conditions which affect less than 1 in 2,000 individuals. Nevertheless, it is estimated that more than 30 million people across the European Union are affected by one of ∼6000-8000 different RDs^1,2^. As 80% of RD are expected to have a genetic aetiology, massively parallel sequencing approaches, in particular exome sequencing (ES), have been widely applied over the last decade to identify variants in DNA that cause RD. However, despite many advances in technology during this period, more than half of all individuals affected by an RD remain without a molecular diagnosis following such analyses, thus extending their diagnostic odyssey. While the accurate detection of single nucleotide variants (SNV) and short (<50nt) insertions and deletions (InDels) from ES data has become relatively robust in recent years^3^, the reliable detection of larger variants, including copy number variants (CNVs), remains a challenge, and it is likely that undetected pathogenic CNVs account for a proportion of undiagnosed individuals.

CNVs comprise losses, which may be heterozygous or homozygous in autosomes, or hemizygous in gonosomes, and gains of genetic material, which we refer to here as *deletions* and *duplications*, respectively. Identification of CNVs from short-read ES data (*i.e.* 100-150nt paired-end reads) is complicated by several factors, the most important of which being that read length is usually shorter than variant length, and that the boundaries of the CNV, referred to as breakpoints, are unlikely to be captured directly by the enrichment targets, since they represent only ∼1-2% of the genome. An exacerbating factor is marked variability in the enrichment process, in which targets for ∼200,000 exons undergo DNA hybridisation and PCR amplification prior to sequencing, both between kits, and between experiments. Many methods have been developed for CNV detection from ES data, most of which use comparison of depth of coverage (DoC) between the observed number of reads covering a particular exon/target in a sample of interest and the normalised coverage for the same exon/target in a large homogeneous reference batch of matched experimental samples^4–9^. For such methods to be successful, the sequencing data needs to be as homogenous as possible, particularly with respect to evenness of coverage^10^, which is the key factor in CNV detection since it directly affects the signal-to-noise ratio.

As reviewed recently in Gordeeva *et al*.^11^, these methods differ from each other primarily in terms of the approach taken for read count normalisation, assumptions regarding read-depth distribution, and the segmentation process, *i.e.* identification of the boundaries of a variant. Despite application of sophisticated normalisation techniques, the correct separation of the signal of true CNVs from background noise remains challenging, particularly for short CNVs that only impact upon one or a few exons. This is illustrated by numerous cross-tool comparisons in which the intersection of CNVs detected by different methods is limited, ranging from ∼1-20% concordance when three or more tools are compared across samples^12–14^. Indeed, a recent benchmarking initiative involving sixteen tools showed that the number of raw CNVs called on a single ES sample ranged from just two to over a thousand^11^, reflecting differing optimisation of algorithms for specificity or sensitivity. Therefore, following identification of a list of potential CNVs, subsequent filtering steps are required, including determining which CNVs are technically valid (*i.e. bona fide* biological events), and whether any of the valid CNVs are of clinical relevance with respect to the phenotype of the affected individual. Hence, both technical expertise and expert clinical knowledge are required if disease-causing CNVs are to be correctly identified. diagnoses.

This complexity may explain why the detection of CNVs has often been omitted from diagnostic ES workflows, with array comparative genome hybridisation (aCGH) continuing to be the preferred method in the clinic over the last decade, despite limitations in its sensitivity and resolution, particularly with respect to short CNVs. However, recent studies have indicated that ES may be a suitable replacement as a first-tier diagnostic test^15–17^, with the added benefit that SNVs and InDels are detected simultaneously.

A key goal of the EU Horizon 2020 Solve-RD project is to raise the diagnostic rate of individuals with an RD for whom ES analysis and variant interpretation have previously been undertaken, but without a conclusive diagnosis having been reached. This is being achieved by undertaking massive pan-European data collation and complete reanalysis from raw data, followed by expert technical and clinical interpretation and validation of variants^18^. The CNV analysis conducted here, was an integral part of a larger re-analysis effort undertaken on the same dataset, covering most other variant types (Laurie *et al.* under review). Here we describe the workflow applied in a comprehensive reanalysis of this heterogeneous sample of ES data from 9,171 individuals pertaining to 5,757 families, including 6,143 individuals affected by an RD, to identify (likely) pathogenic CNVs. The ES data had been generated using 28 different enrichment kits in multiple sequencing centres. Hence, to maximise accuracy and sensitivity of CNV detection we applied three different algorithms, ClinCNV, Conifer, and ExomeDepth, and analysed experiments in 28 different batches, comprising data generated using the same enrichment kit. We filtered the raw call set, initially consisting of over two million CNV calls (average of ∼300 per individual), to a manageable number of 0-2 potentially pathogenic rare CNVs per affected individual requiring interpretation by the clinical experts who has submitted the cases to Solve-RD. This extensive endeavour has led to the closure of many diagnostic odysseys, some of which had been ongoing for decades, of which we provide some illustrative examples.

## Methods

### Data Collation

The ES data reanalysed here comprises previously inconclusive ES experiments submitted for reanalysis as part of the Solve-RD project by 42 different research groups based in twelve countries across Europe, and Canada (range of 1-2,111 experiments submitted per group). Each experiment was submitted via one of four European Reference Networks (ERN) partnering in Solve-RD, each focusing on a particular group of RD: EURO-NMD (rare neuromuscular diseases); GENTURIS (rare genetic tumour risk syndromes); ITHACA (rare malformation syndromes, intellectual and other neurodevelopmental disorders); RND (rare neurological diseases).

A total of 9,351 ES experiments from 9,314 individuals (6,224 affected individuals and 3,090 unaffected relatives) were initially submitted for reanalysis. After the removal of samples sequenced with enrichment kits for which the available control cohort was not large enough to allow accurate CNV identification, data from 9,171 individuals from 5,757 families was analysed (see Technical Results). While 1,320 of 1,788 (74%) families from ITHACA were composed of parent-child trios, facilitating identification of *de novo* mutations, only 239 of the remaining 3,969 (6%) probands from other ERNs were trios. ES had been performed using 28 different enrichment kits (range of 4-2,078 experiments per kit), and each of the forty-two research groups had followed their own DNA library preparation, target enrichment, and short-read sequencing protocol in their local labs, or via external DNA sequencing providers. Furthermore, each group had previously undertaken their own historic analysis and interpretation of the resulting ES data to identify disease-causing variants, which has proven inconclusive. The date at which the initial ES analysis and interpretation had been undertaken ranged from six months to eight years prior to the experimental data being submitted to Solve-RD for reanalysis, however this information was not collected systematically for individual data sets.

In addition to sequencing data, a phenotypic description for each affected individual was recorded in the PhenoStore module of the RD-Connect GPAP^19^, consisting of a minimum of five Human Phenotype Ontology terms (HPO^20^) wherever possible, and disease classification using Orphanet Rare Disease Ontology (ORDO) ORPHA codes (http://www.orphadata.org/cgi-bin/index.php), and/or OMIM identifiers^21^ (https://www.omim.org/) where appropriate, together with family pedigrees. A detailed description of this data set can be found in Laurie et al, 2023 (under review).

### CNV Identification

Raw ES data was realigned to the hs37d5 reference genome^22^, using BWA-MEM^23^, as described in the Supplementary Materials. With the goal of maximising the probability of detecting potentially disease-causing CNVs, three different algorithms which identify CNVs based on DoC were applied. Two of these, Conifer^4^, and ExomeDepth^6^, have been widely applied to ES data with success previously, while the third, ClinCNV, was developed recently by a Solve-RD partner^24^. Each of these tools offers the practical advantage of separating the DoC calculation for each individual experiment from the CNV calling step, and thus CNVs were subsequently called in batches by enrichment kit. Furthermore, each algorithm provides an estimate of the likelihood that calls produced are biologically real, and the most likely false positive calls were excluded based upon these metrics. As primary filters, in the case of Conifer a value in excess of +/-1.75 SV-RPKM was required for a CNV call to be taken forward for biological interpretation, while for ExomeDepth a Bayes Factor (BF) greater than fifteen was required, and for ClinCNV a minimum log-likelihood estimation of twenty was applied (see Supplementary Methods for further detail).

### Call Filtering and Visualisation

As the focus of Solve-RD is diagnosing RD cases, through the identification of rare variants that are potentially disease-causing, any apparent CNV call observed in a region where more than 1% of individuals in the whole sample had a similar type of call (*i.e.* a deletion or duplication) were discarded as being too common to be clinically relevant with respect to RD. Furthermore, CNVs returned for interpretation by clinical experts were restricted to those which overlapped with at least part of a gene in a predefined list of curated genes of interest provided by the respective ERN: EURO-NMD (n=615), GENTURIS (230), ITHACA (1,944), RND (1,820). The full list of ERN curated genes is provided in **Supplementary Table 1** and details as to how these lists were determined in Laurie et al, 2023 (under review). Potential CNVs of interest were subsequently categorised into six non-redundant classes to aid interpretation: Long CNVs (>500kb in length) ; Homozygous deletions; Heterozygous CNVs affecting genes known to cause disorders with an autosomal dominant mode of inheritance; Regions with apparent copy numbers of four or more; Gonosomal CNVs ; Potential compound-heterozygous *double-hits* in the form of a CNV affecting the second allele of a gene in which biallelic variants are known to be disease-causing, and in which a potentially pathogenic SNV has been previously identified in Solve-RD.

To provide support for interpretation of the technical validity of CNV calls, images of regions containing CNV calls were generated automatically using the Integrative Genomics Viewer (IGV)^25^. A variety of custom tracks, including call tracks for each of the three algorithms, BAM DoC, and gene tracks for ERN genes of interest were incorporated, among others (see Supplementary Methods).

### Clinical Interpretation

Further annotations to aid interpretation (**Supplementary Table 2**) were added to the results using AnnotSV^26^ (Version 3.0.7), and fully annotated CNV call sets generated for all tools together with accompanying customised IGV visualisations were distributed to clinical experts in each ERN for diagnostic interpretation. Each ERN prioritised calls for further investigation based on their expert knowledge of underlying disease mechanisms in their respective patients. Many CNV calls could be rapidly discarded based upon a lack of match between the gene potentially affected and the phenotype of the affected individual, and/or segregation patterns within the family. Others were rejected when visual inspection of the IGV tracks indicated that they were likely false-positive calls, and thus unlikely to be *bona fide* biological events. Where deemed necessary and when feasible, CNVs believed to be diagnostically relevant were validated at local centres using orthologous approaches. The final decision as to whether a CNV was determined to be pathogenic or not was taken by the respective clinical experts from the ERN (see Supplementary Methods for further details).

## Results

### Technical Results

Prior to the initiation of CNV calling, a minimal quality control was undertaken, which took the form of requiring that data from each submitted family included at least one affected individual with accompanying HPO terms. Furthermore, following alignment of sequencing reads, it was required that at least 70% of the target region of the enrichment kit had a DoC of ten reads. After removal of 143 experiments which did not meet these criteria, CNV calling was undertaken on data from a total of 9,171 individuals from 5,757 families, of whom 6,143 had a rare condition. Initial investigations indicated the presence of a large variance in sequencing depth both within and between the twenty-eight enrichment kit batches, reflecting the heterogeneity of the sequencing data submitted to Solve-RD (**Supplementary Figure 1**).

Following identification and removal of likely false positive calls based upon tool-specific QC metrics, the removal of commonly observed events, and restriction to events overlapping genes in the custom gene lists from the corresponding ERN, a total of 7,849 calls in 3,436 affected individuals from 3,300 families remained for interpretation (**Table1**). The number of probands with at least one CNV call to be interpreted by clinical specialists from the ERN ranged from 113 for GENTURIS (33% of families), to 1,239 for ITHACA (69% of families) (**Supplementary Table 3**). No CNV of interest was detected in 2,707 affected individuals from the remaining 2,457 families. In addition, a further 393 pairs of potential CNV-SNV *double-hit* compound heterozygous variants in 226 affected individuals were returned to clinical experts for interpretation. Overall, a mean of 1.3 CNVs per proband were returned for interpretation. However, as CNVs of potential interest were only identified in 55% of probands, this equated to 2.4 variants per proband that required interpretation.

The total number of CNV calls in affected individuals returned for interpretation was highest for ExomeDepth (n=4,205), while ClinCNV called about two-thirds of this number (2,782), and Conifer approximately one-fifth (862), reflecting different predilections of the underlying algorithms with respect to sensitivity and specificity of CNV detection. While Conifer and ExomeDepth showed a significant bias towards calling duplications, the reverse pattern was observed for ClinCNV, which identified more deletions (p<0.00001 in all cases, Fisher exact test; **Supplementary Table 4).** We assessed the distribution of the length of CNVs returned for interpretation as identified by each too. Notably, the average length of CNVs detected by Conifer was approximately an order of magnitude larger than that of ExomeDepth, which in turn was longer than that of ClinCNV. This pattern held for both duplications and deletions, and again reflects differences in the way the tools identify and segment CNVs (**Figure 1, Supplementary Table 5**).

**Figure 1.**
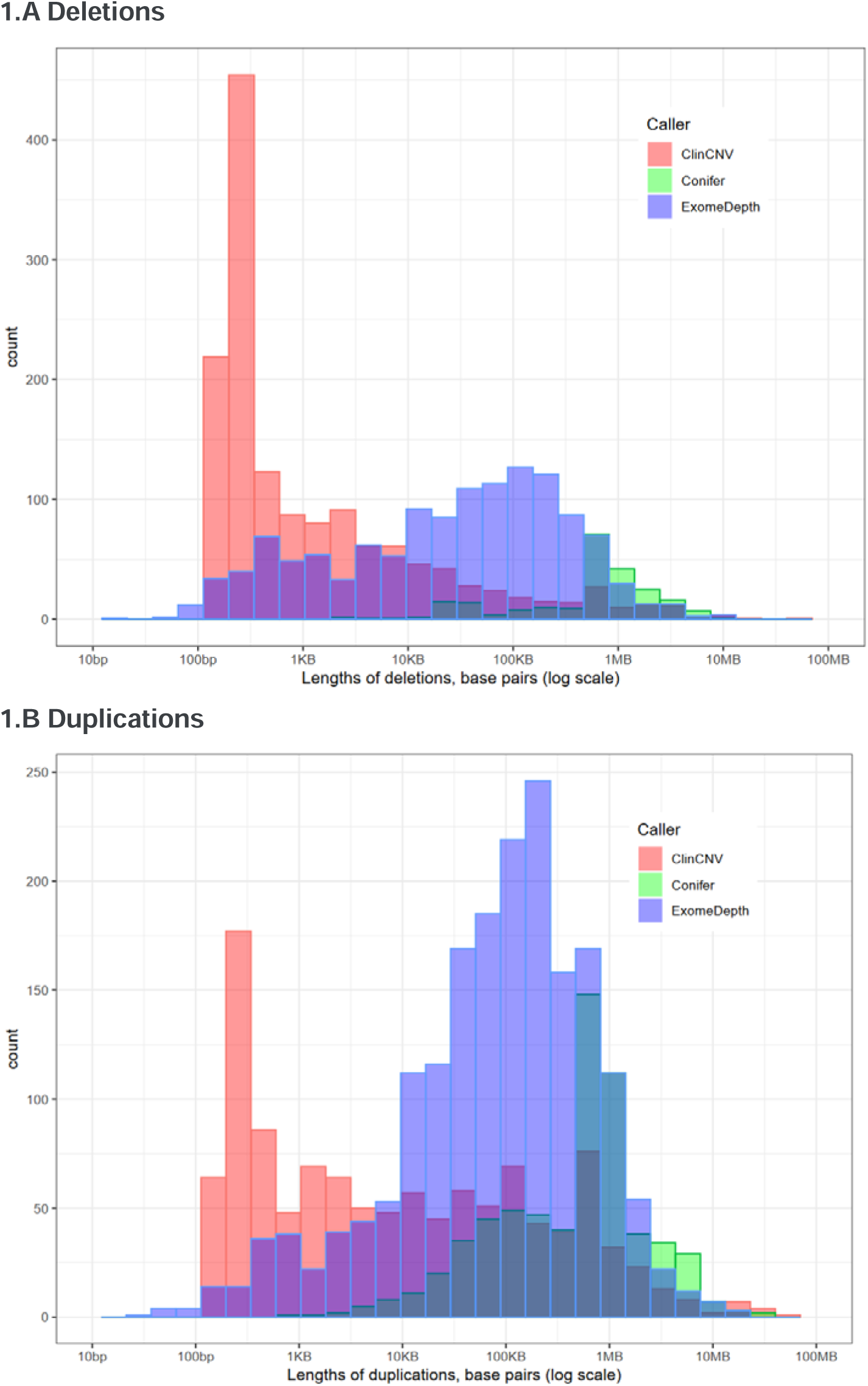
Distribution of lengths of 7,849 CNV calls detected in 3,436 affected individuals, separated into deletions (Panel A) and duplications (Panel B). The x-axis represents the length of calls identified (log_10_ scale), and the y-axis the number of events observed. ClinCNV is represented in red. Note that the y-axis scale is different in 1.A from 1.B

### Diagnostic Results

Following expert interpretation, 105 potentially pathogenic CNVs of interest in 103 affected probands were identified, of which 52 have been confirmed as disease-causing in 51 individuals (**Table 2**). The disease-causing CNVs included three “double-hit” instances where an SNV and CNV affecting different alleles of the same gene were identified, resulting in a compound heterozygous diagnosis, and one instance where two CNVs affecting different genes provided a dual genetic diagnosis for a complex phenotype. A further 25 CNVs are regarded as pathogenic by the clinical experts, but not sufficient, to explain the full phenotype observed in the affected individual, including seven complete gonosomal aneuploidies (“Partially Explanatory” in **Tables 2 and 3**). A further 28 potentially pathogenic CNVs were identified for which further validation is not logistically possible due to lack of access to DNA and/or the patient (referred to as candidates below). While 81% (42 of 52) of confirmed disease-causing CNVs are deletions, only 39% (7 of 18) of the partially explanatory pathogenic CNVs are deletions, even when disregarding the gonosomal duplications. Of the 28 candidate CNVs 57% (16) are deletions (**Figure 2**, **Table 2**).

**Figure 2.**
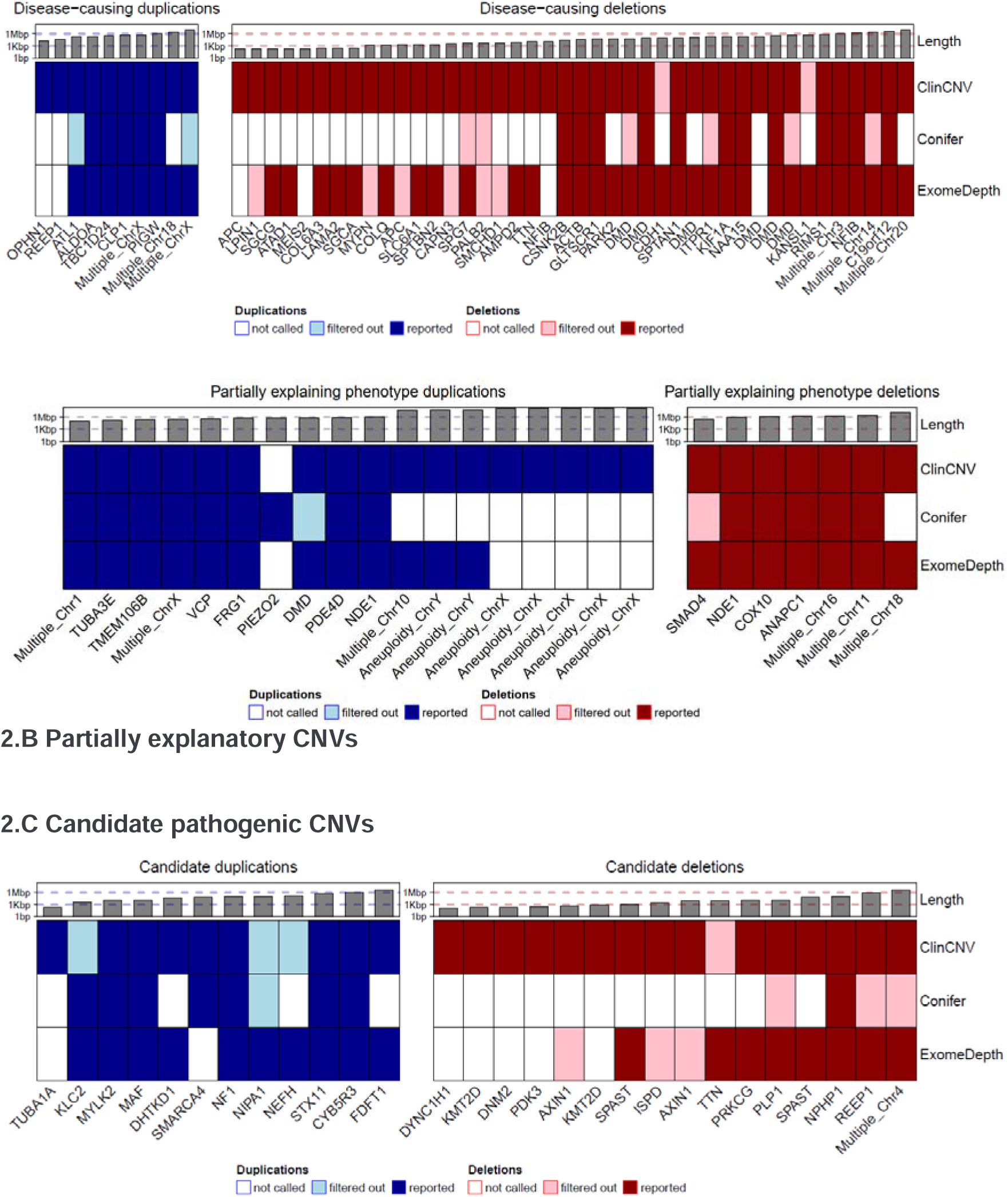
Heat maps illustrating the length of confirmed disease-causing CNVs (Panel A), partially explanatory disease-causing CNVs (Panel B) and candidate disease-causing CNVs (Panel C) identified in this study. Duplications are shown in blue, and deletions in red. Cyan and pink, represent duplication and deletion calls, respectively, which were not reported back for interpretation due to being QC filtered in the workflow for the respective tool. The approximate length of the event is indicated in the top layer using a log_10_ scale. The affected gene is indicated along the bottom. Where more than one gene was unaffected, it is shown as multiple, with the affected chromosome indicated.

Of the 77 confirmed pathogenic CNVs, 40 (52%) were initially identified by all three callers (**Figure 2**, **Table 2)**. However, in the case of ten of the 40, the Conifer call was subsequently discarded due to it being within the applied SV-RPKM threshold, and one of the ten was also discarded by the ExomeDepth workflow due to a low BF. Of the remaining 37 pathogenic CNVs, 36 (97%) were identified by ClinCNV, two of which subsequently failed ClinCNV quality control thresholds, while 25 (68%) were identified by ExomeDepth, five of which were subsequently discarded due to a low BF. Interestingly one of the 37, a duplication in *PIEZO2* was identified by Conifer alone.

### Examples of successful new diagnoses

Below we provide an example of an RD case from each of the four ERN partners in Solve-RD solved through the analysis of CNVs undertaken here.

### ERN EURO-NMD

This male in his thirties first came to clinical attention in his adolescence, affected by poor balance, recurrent falls, and difficulty rising from the floor. Prior to this he had been able to run and play sports normally. His symptoms worsened slowly over time, and he is currently unable to walk or stand without assistance. He also has mild facial weakness and mildly elevated serum creatine kinase. His family history is negative, having several unaffected siblings. Muscle biopsy showed clear features of muscular dystrophy, and immunohistochemical analysis suggested reduced expression of dystrophin. Exome sequencing was initially undertaken in 2017, but no diagnosis was reached at that point.

As a result of reanalysis of the ES data undertaken here, a three-exon deletion affecting exons 45 through 47 of the *DMD* gene was detected by both ExomeDepth and ClinCNV, consistent with the suspected diagnosis of Becker Muscular Dystrophy. This hemizygous deletion was subsequently confirmed using Multiplex Ligation-dependent Probe Amplification (MLPA). Confirmation of the molecular diagnosis in this individual has enabled enhanced genetic counselling, as any future daughter he may have would be an obligate, and possibly manifesting, carrier of the CNV, thus requiring clinical management.

### ERN GENTURIS

This family first came to clinical attention in 2003, meeting the criteria for hereditary diffuse gastric cancer (HDGC)^27^, as several family members had developed diffuse gastric cancers prior to 30 years of age. HDGC typically results from *CDH1* loss of function^28,29^. However, Sanger sequencing of *CDH1* performed proved negative, as did subsequent investigation in the form of MLPA, and ES, at which point no potentially explanatory SNVs, InDels, or CNVs were identified in *CDH1*, nor other candidate genes^30^.

Following these negative findings, the ES data was submitted to Solve-RD for two affected, and four unaffected siblings. The comprehensive reanalysis of the ES data resulted in the identification of a ∼116kb heterozygous deletion impacting half of the *CDH1* gene (from intron 7 forwards) and the start of the downstream gene *TANGO6* (as far as intron 14) (g:16:68846036-68964198del) in four of the six siblings (**Supplementary Figure 2**). The CNV was detected by both ClinCNV and ExomeDepth and further supported by split-reads and abnormally paired reads observed in data from one of the affected individuals. Visualisation in IGV, and subsequent MLPA, validated this large event. Of note, one of the unaffected siblings, a female carrier in her forties, has not developed gastric cancer to date, in accordance with previously reported incomplete penetrance among *CDH1* mutation carriers^31^. Another of the unaffected siblings was a carrier but never developed gastric cancer as a result of having undergone prophylactic total gastrectomy due to the high incidence of cancer in the family. The remaining unaffected siblings were found not to harbour the deletion, but unfortunately both have also already undergone prophylactic gastrectomy. Nevertheless, as a result of their inclusion in Solve-RD, the family has since been recontacted and enrolled in a clinical pathway of care, and their twenty-year diagnostic odyssey has now come to an end. Importantly, targeted genetic testing has now been made available to their offspring to avoid unnecessary prophylactic gastrectomy in subsequent generations. The functional analysis and clinical implications of this CNV are described in more detail in São José et al.^32^.

### ERN ITHACA

This girl was first referred to paediatric neurology in her first year of life, presenting with generalised tonic-clonic seizures. During her infancy mild global developmental delay became evident, with delays in speech and language acquirement and in gross-motor skill acquisition Seizures were controlled with lamotrigine monotherapy, which could be discontinued during childhood following prolonged seizure-free periods. Apart from polyhydramnios, pregnancy and delivery were uncomplicated. Medical history comprised constipation and eczema, and family history was unremarkable. Physical examination revealed no additional phenotypic features *i.e.* no congenital anomalies, no facial dysmorphisms, and no growth abnormalities. Investigations, including cerebral MRI and general metabolic screening. Singleton ES was performed, followed by trio ES which revealed a heterozygous *de novo* SNV of uncertain significance (VUS) in *STIP1* (*STIP1*; Chr11(GRCh37):g.63961718C>T; NM_001282652.1:c.418C>T; p.(Arg140*)). Within this diagnostic trajectory, no analysis dedicated to CNV detection was performed.

The systematic reanalysis of ES data reported here led to the identification of a heterozygous 27kb deletion on chromosome 6p21 (chr6:31630124-31657924-DEL) in the proband. This deletion was detected by all three tools, and visual inspection of sequence alignment files in IGV clearly indicated the presence of the variant in the affected daughter, and its absence in both parents, thus confirming that it is a *de novo* deletion. The deletion fully removes *CSNK2B, LY6G5B* and *LY6G5C*, and its breakpoints affect *GPANK1* and *ABHD16A*. *GPANK1, LY6G5B* and *LY6G5C* currently have no disease association, and while *ABHD16A* is associated with autosomal recessive spastic paraplegia-86 (MIM#619735), there is no apparent second hit in *ABHD16A*, and the phenotype of the proband does not comprise spastic paraplegia. *CSNK2B,* on the other hand, has recently been shown to be associated with autosomal dominant Poirier-Bienvenu neurodevelopmental syndrome (POBINDS; MIM#618732), in which truncating variants in *CSNK2B* result in haploinsufficiency, leading to early-onset seizures and highly variable impairments of intellectual functioning^33–35^. As the *de novo* deletion observed in this proband results in haploinsufficiency of *CSNK2B,* and her phenotypic descriptions fits within the *CSNK2B*-associated phenotypic spectrum, this 27kb deletion on chromosome 6p21 is regarded as explanatory for her rare condition, thus ending a seven-year diagnostic odyssey for this family.

### ERN RND

This teenage female was first evaluated in paediatric neurology as a child, presenting with global developmental delay, and behavioural and learning problems. Retrospectively, the first symptoms had become apparent in her infancy, consisting of mild delayed development of fine and gross motor skills. Additionally, she had delays in language and speech development, and was diagnosed with attention deficit disorder, for which she is being treated with methylphenidate and responding well. No obvious dysmorphic features were observed upon physical examination, but mild hypertonia of the triceps surae, hyperreflexia, kinetic tremor, mirror hand movement, and a tiptoeing gait were observed. Subsequent cerebral MRI showed ventriculomegaly, corpus callosum hypoplasia, prominent cerebellar folia, and thin middle cerebellar peduncles. Genetic testing, consisting of aCGH (median resolution 180kb), targeted testing for Fragile X syndrome, and ES did not pinpoint a molecular cause.

Systematic reanalysis of the ES data undertaken here led to the identification of a heterozygous deletion of ∼200kb at chromosome 4q31.1: Chr4(GRCh37):g.140187686-140394334del, encompassing part of the *MGARP* gene (not known to be associated with disease), and the entire *NAA15* gene, which encodes the catalytic subunit in the N-terminal acetyltransferase A complex (MIM: 608000). The deletion was identified by all three tools, and subsequently validated using high resolution aCGH (median resolution 60kb). Following review of the prior results, the absence of recall of the variant in the initial aCGH analysis was attributed to its limited resolution. The patient’s mother, who had had similar learning problems and has mild cognitive disability, was subsequently also found to be positive for the deletion. No further family testing was possible. Echocardiography was normal in both cases. Loss-of-function variants in *NAA15* and heterozygous deletion of this gene and nearby genes are associated with ‘Intellectual developmental disorder, autosomal dominant 50, with behavioural abnormalities’ (MIM: 617787)^36,37^. This disorder has the features of a wide spectrum of neurodevelopmental severity and variable association of congenital anomalies, thus confirming that the observed CNV was causative in this case and ending this family’s seven-year diagnostic odyssey.

## Discussion

Rigorous detection of CNVs from ES requires sequencing data that has been generated as uniformly as possible, in order that the test experiment can be compared against a similarly generated batch of matched control samples. However, the ES data submitted to Solve-RD had been generated using twenty-eight different enrichment kits and sequenced with different short-read technologies to different depths of coverage, in multiple sequencing centres across Europe. Hence the primary challenge encountered during this analysis was data heterogeneity. Similarly, from the perspective of diagnosis, it is essential to have a clear clinical description of the affected individual to be able to determine in which genes, variants, if encountered, may explain the observed phenotype. This was achieved here firstly through use of the HPO ontology to capture a deep phenotypic description of affected individuals from the referring clinicians, and secondly using the curated set of genes of interest provided by each ERN. Together these significantly reduced the search space for potentially disease-causing CNVs.

The interpretation of raw CNV calls is challenging due to the initial high number of calls most tools report. We applied a robust filtering strategy to remove calls that were clearly unlikely to be of relevance for RD and benefited from the curated lists of genes of interest provided by each ERN. Nevertheless, visual inspection of the affected region using IGV was key for assessing the technical validity of remaining calls, prior to, or in parallel with, their biological interpretation. It is likely that this is an aspect where an AI-based tool for automated IGV-image analysis would be of significant benefit, potentially saving many hours of human expert-review time. The clinical researchers representing each ERN applied their own prioritisation strategy when interpreting CNV calls, according to the specific pathologic and phenotypic characteristics of their patients.

When used as a first-tier analysis, CNV detection from ES has been reported to result in diagnostic yields as high as 7-19%^38–40^, whereas yield The overall rate of novel diagnoses reached was 0.9%, ranging from 0.6% for RND and 0.9% for ITHACA to 1.2% for GENTURIS and EURO-NMD. Notably nine of the sixteen CNVs established as being disease-causing in ITHACA cases could be confirmed as *de novo* mutations due to ES data being available from the proband’s parents. While our values are lower than those of prior reports, where yield from reanalyses efforts, have resulted in increases in diagnostic yield with respect to CNVs in the range of 1.6-2.0%^41–43^, in those studies the prior CNV analyses had largely consisted of only chromosomal microarray (CMA) analyses, which lack sensitivity for short CNV events which were hence identified in the subsequent ES-based CNV analyses. Our results reflect several factors: the likelihood that detailed CNV analysis of the ES had been undertaken prior to submission to Solve-RD; the role that CNVs are likely to have in the respective class of disease; the time passed since the initial analysis, which would affect the number of genes known to be associated with a particular class of disease. Interestingly, the number of genes of interest in each of the custom ERN gene lists does not appear to be a factor given that GENTURIS had by far the shortest list, and RND and ITHACA the longest.

There was a clear bias towards deletions *vis-à-vis* duplications being identified as pathogenic with 49 of 77 (64%) confirmed pathogenic CNVs being deletions, and 42 of 52 (81%) disease-causing CNVs. This reflects the facts that duplications are more challenging to detect, and even when detected by ES, it is invariably unclear as to whether they are tandem duplications, possibly inverted, or inserted elsewhere in the genome, each of these scenarios being likely to result in a different biological consequence, making interpretation challenging. Furthermore, long duplications appear to be under less evolutionary constraint than similarly sized deletions^44^, suggesting that they are less likely to results in disease. Accordingly, the ACMG guidelines for the interpretation of constitutional CNVs^45^, require more supporting evidence for a duplication to be confirmed as pathogenic than is required for a deletion.

It is noteworthy that, in comparison with the other two tools, Conifer called very few CNVs under 20kb in length, and indeed failed to successfully identify 18 of 20 deletions <20kb that were determined to be disease-causing, and the remaining two fell below the calling threshold. Notably, Conifer also failed to identify duplications over 1Mb in length, including seven sex-chromosome aneuploidies. It is this failure at the two extremes of CNV length that largely contribute to the inferior performance of Conifer. It should also be highlighted that we required a Z-score in excess of +/-1.75 for a CNV called by Conifer to be returned for interpretation, whereas had we used +/-1.5, Conifer would have successfully identified a further eight events of the disease-causing CNVs, all but two of which were over 20kb in length. ClinCNV performed best of the three callers with this highly heterogeneous dataset, which is likely due to its more adaptive DoC calculation whereby it subsegments target regions into 120bp tiles, significantly improving resolution, particularly for short CNVs, most of which were also detected by ExomeDepth but some fell below the minimal calling threshold.

In addition to cases of *de novo* dominant inheritance resolved by an individual CNV, we also identified eight cases where an SNV and CNV were affecting different alleles of the same gene potentially forming a disease-causing compound heterozygote. Two of these have been confirmed as being explanatory for the individuals’ conditions, with the remaining six requiring further validation. These findings underline the importance of having all data relevant to the interpretation of an affected individual’s condition readily at hand, as had the SNV and CNV analyses been undertaken independently, these individuals would have been unlikely to have received a diagnosis. Furthermore, in one affected individual, we identified two pathogenic CNVs affecting different genes, each of which explain unique features of the individual’s complex phenotype, *i.e.* a dual diagnosis^46^. We are confident that many of the CNVs that we currently classify as candidates are likely pathogenic in the affected individuals, but complete follow-up has not yet been possible. The complete expert-curated dataset of deletions and duplications, together with the detailed phenotypes and pedigrees, and the aligned sequence files (CRAMs) are available to the entire RD community via the European Genome-Phenome Archive (EGA)^47^, allowing for new discoveries.

There are many reasons why a pathogenic CNV identified here may not have been found in prior analysis of the ES data. Firstly, there may have been no attempt to identify CNVs by the respective clinical research team, due to a lack of resources or expertise. However, we know that some form of prior CNV analysis had been undertaken for the majority of affected individuals analysed here. Secondly, the tool(s) applied previously for CNV detection may not have identified the relevant CNV, or though identified, it may have been discarded due to local quality control parameters applied *e.g.* approximately 10% of all the experiments submitted to Solve-RD were of sufficiently poor quality such that one of the centres involved in the reanalysis undertaken here would have routinely QC-failed the sample in their diagnostic workflow and thus not attempted to identify CNVs. Thirdly, while the CNV may have been identified, there may not have been any known association between the affected gene(s) and the clinical presentation of the patient at the time of the initial analysis, resulting in, at best, classification of the CNV as a variant of uncertain significance (VUS), and the individual remaining undiagnosed.

We would emphasise that any observations of potential tendencies in the results presented here must be interpreted prudently since this was an extremely heterogeneous dataset both in terms of the breadth and the quality of the data, and in terms of the time and expertise that had been applied to the interpretation of the ES data in analyses undertaken prior to submission to Solve-RD. As we gather more information about the role of CNVs in RD through projects that share data widely such as Solve-RD, hopefully the accuracy of CNV detection will improve, and the entire process of identification and interpretation of this important class of variants, from sequencing data to identification of pathogenic variants can be automated, resulting in families affected by RD receiving a diagnosis sooner rather than later.

### Limitations of this work

The work presented here has several clear limitations *vis-a-vis* reaching a diagnosis for individuals affected by an RD. Firstly, given that the data was from ES, and that we only considered events affecting one of between 230 and 1,944 genes of interest identified by each of the ERNs, we will obviously miss any non-exonic events, or CNVs affecting genes not in the list of genes of interest. However, undertaking this work without using gene lists would result in a currently insurmountable load of data for interpretation, and novel gene discovery was not the goal of the work undertaken here. However, such discoveries are enabled by the sharing of data with the wider RD community vie the EGA, which we hope will enable more cases to be solved. Different approaches in interpretation undertaken by the ERN experts may have resulted in some biologically relevant events being discarded as uninteresting, which may be particularly true for duplications, for which evidence of biological relevance in RD is currently relatively scarce. It is also possible that application of other tools designed to find CNVs affecting only single exons, such as VarGenius-HZD^48^, may have allowed the identification of shorter events missed by the tools applied. With the future adoption of long-read genome sequencing technologies such as those provided by Oxford Nanopore Technologies and Pacific Biosciences, it is likely that the accuracy of CNV detection, and hence ease of interpretation, will improve markedly.

Despite these limitations, we have successfully provided diagnoses to at least 51 families who had previously undergone extensive genetic testing and in many cases multiple hospital visits over many years, some even decades, without having been provided with a diagnosis. Within the larger Solve-RD reanalysis of all variant types, these 51 CNVs were the second most common type of disease-causing variant identified, after SNVs/InDels, contributing to ∼9% of the successful diagnoses Laurie et al. 2023 (under review). The ending of a diagnostic odyssey has many benefits to patients and their families, beyond changes in medical management and genetic counselling of relatives. It also allows better understanding of disease progression, access to disease-specific online communities, and psychological closure, amongst other benefits^49^. The work undertaken here indicates the value of comprehensive (re)-analysis of copy number variants in undiagnosed RD cases, even from historic ES data, and has resulted in patients and their families being given an accurate diagnosis, finally ending their diagnostic odysseys.

### Recommendations

Based upon our findings, we suggest the following recommendations for future (re)-analyses of ES data with respect to identification of disease-causing CNVs.

1) Know your enrichment kit. Investigate how well, and how evenly, does it capture your genes of interest.
2) Choose your tools wisely. While Conifer has been shown to work with homogenous datasets e.g. thousands of ES datasets generated using the same kit, in the same sequencing centre, it does not perform with the heterogeneous dataset analysed here. Furthermore, it identified very few CNVs <20kb in length, missing many disease-causing variants.
3) Identifying regions that are commonly copy-number variant. In this way any CNVs observed in such regions can be excluded from being potentially disease-causing.
4) Use an *in silico* candidate gene list when possible. This will greatly accelerate the process of interpretation. If the list is very short, then any signal of a CNV in a gene of interest should be examined further, since the sensitivity of tools remains low, and the prior probability of the gene being variant is high. However, as lists grow longer, this probability reduces, and calls will have to be filtered by quality thresholds.
5) Visualisation of CNV calls using a tool such as IGV is essential to assure that they are likely to be real biological events, prior to expending time and effort on further interpretation, investigation, and/or confirmation using orthologous techniques.

## Data Availability

All data produced in the present study will be deposited at the EGA, Hinxton, Cambridge, in due course.

## Data availability and Ethics Statement

Data will be deposited at EGA. Accession numbers to be provided. The family (FAM) and participant (P) identifiers used in this manuscript are pseudonymised and known only to the researchers involved In Solve-RD. The Ethics committee of the Eberhard Karl University of Tubingen gave ethical approval for this work.

## Corporate author list

### Solve-RD consortium

**EKUT:** Olaf Riess^1, 2^, Tobias B. Haack^1^, Holm Graessner^1, 2^, Birte Zurek^1, 2^, Kornelia Ellwanger^1, 2^, Stephan Ossowski^1, 3^, German Demidov^1^, Marc Sturm^1^, Julia M. Schulze-Hentrich^1^, Rebecca Schüle^1, 2^, Jishu Xu^4, 5^, Christoph Kessler^4, 5^, Melanie Kellner^4, 5^, Matthis Synofzik^4, 5^, Carlo Wilke^4, 5^, Andreas Traschütz^4, 5^, Ludger Schöls^4, 5^, Holger Hengel^4, 5^, Holger Lerche^1^, Josua Kegele^6^, Peter Heutink^4, 5^

**RUMC:** Han Brunner^7–9^, Hans Scheffer^7, 8^, Nicoline Hoogerbrugge^7, 10^, Alexander Hoischen^7, 10, 11^, Peter A.C. ’t Hoen ^10, 12^, Lisenka E.L.M. Vissers^7, 8^, Christian Gilissen^7, 10^, Wouter Steyaert^7, 10^, Karolis Sablauskas^7^, Richarda M. de Voer^7, 10^, Erik-Jan Kamsteeg^7^, Bart van de Warrenburg^8, 13^, Nienke van Os^8, 13^, Iris te Paske^7, 10^, Erik Janssen^7, 10^, Elke de Boer^7, 8^,Marloes Steehouwer^7^, Burcu Yaldiz^7^, Tjitske Kleefstra^7, 8^

**University of Leicester:** Anthony J. Brookes^14^, Colin Veal^14^, Spencer Gibson^14^, Vatsalya Maddi^14^, Mehdi Mehtarizadeh^14^, Umar Riaz^14^, Greg Warren^14^, Farid Yavari Dizjikan^14^, Thomas Shorter^14^

**UNEW:** Ana Töpf^15^, Volker Straub^15^, Chiara Marini Bettolo^15^, Jordi Diaz Manera^15^, Sophie Hambleton^16^, Karin Engelhardt^16^

**MUH:** Jill Clayton-Smith^17, 18^, Siddharth Banka^17, 18^, Elizabeth Alexander^18^, Adam Jackson^17, 18^

**DIJON:** Laurence Faivre^19-23^, Christel Thauvin^19-23^, Antonio Vitobello^21^, Anne-Sophie Denommé-Pichon^21^, Yannis Duffourd^21, 22^, Ange-Line Bruel^21^, Christine Peyron^24, 25^, Aurore Pélissier^24, 25^

**CNAG-CRG:** Sergi Beltran^26, 27^, Ivo Glynne Gut^26, 27^, Steven Laurie^26^, Davide Piscia^26^, Leslie Matalonga^26^, Anastasios Papakonstantinou^26^, Gemma Bullich^26^, Alberto Corvo^26^, Marcos Fernandez-Callejo^26^, Carles Hernández^26^, Daniel Picó^26^, Ida Paramonov^26^, Hanns Lochmüller^26^

**EURORDIS:** Gulcin Gumus^28^, Virginie Bros-Facer^29^

**INSERM-Orphanet:** Ana Rath^30^, Marc Hanauer^30^, David Lagorce^30^,Oscar Hongnat^30^,Maroua Chahdil^30^,Emeline Lebreton^30^

**INSERM-ICM:** Giovanni Stevanin^31–35^, Alexandra Durr^31–34, 36^, Claire-Sophie Davoine^31–35^, Léna Guillot-Noel^31–35^, Anna Heinzmann ^31–34, 37^, Giulia Coarelli^31–34, 37^

**INSERM-CRM:** Gisèle Bonne^38^, Teresinha Evangelista^38^, Valérie Allamand^38^, Isabelle Nelson^38^, Rabah Ben Yaou^38–40^, Corinne Metay^38, 41^, Bruno Eymard^38, 39^, Enzo Cohen^38^, Antonio Atalaia^38^, Tanya Stojkovic^38, 39^

**Univerzita Karlova:** Milan Macek Jr.^42^, Marek Turnovec^42^, Dana Thomasová^42^, Radka Pourová Kremliková^42^, Vera Franková^42^, Markéta Havlovicová^42^, Petra Lišková^43, 44^, Pavla Doležalová^45^

**EMBL-EBI:** Helen Parkinson^46^, Thomas Keane^46^, Mallory Freeberg^46^, Coline Thomas^46^, Dylan Spalding^46^ Jackson Laboratory: Peter Robinson^47^, Daniel Danis^47^

**KCL:** Glenn Robert^48^, Alessia Costa^49^, Christine Patch^49, 50^

**UCL-IoN:** Mike Hanna^51^, Henry Houlden^52^, Mary Reilly^51^, Jana Vandrovcova^52^, Stephanie Efthymiou^52^, Heba Morsy^52^, Elisa Cali^52^, Francesca Magrinelli^53^, Sanjay M. Sisodiya^54^, Jonathan Rohrer^55^ UCL-ICH, Francesco Muntoni^56, 57^, Irina Zaharieva^56^, Anna Sarkozy^56^

**Universiteit Antwerpen:** Vincent Timmerman^58, 59^, Jonathan Baets^60, 61^, Geert de Vries^59, 60^, Jonathan De Winter^59–61^, Danique Beijer^58–60^, Peter de Jonghe^59, 61^, Liedewei Van de Vondel^58–60^, Willem De Ridder^59–61^, Sarah Weckhuysen^60, 62^

**Uni Naples/Telethon UDP:** Vincenzo Nigro^63, 64^, Margherita Mutarelli^64, 65^, Manuela Morleo^64^, Michele Pinelli^64^, Alessandra Varavallo^64^, Sandro Banfi^63, 64^, Annalaura Torella^63^, Francesco Musacchia^63, 64^, Giulio Piluso^63^

**UNIFE:** Alessandra Ferlini^66^, Rita Selvatici^66^, Francesca Gualandi^66^, Stefania Bigoni^66^, Rachele Rossi^66^, Marcella Neri^66^

**UKB:** Stefan Aretz^67, 68^, Isabel Spier^67, 68^, Anna Katharina Sommer^67^, Sophia Peters^67^

**IPATIMUP:** Carla Oliveira^69–71^, Jose Garcia-Pelaez^69, 70, 72^, Rita Barbosa**-**Matos^69, 70, 73^, Celina São José^69, 70, 72^, Marta Ferreira^69, 70, 74^, Irene Gullo^69–71, 75^, Susana Fernandes^76^, Luzia Garrido^75^, Pedro Ferreira^69, 70, 77^, Fátima _Carneiro_69-71, 75

**UMCG:** Morris A Swertz^78^, Lennart Johansson^78^, Joeri K van der Velde^78^, Gerben van der Vries^78^, Pieter B Neerincx^78^, David Ruvolo^78^, Kristin M Abbott^79^, Wilhemina S Kerstjens Frederikse^79, 80^, Eveline Zonneveld-Huijssoon^79, 81^, Dieuwke Roelofs-Prins^78^, Marielle van Gijn^79, 81^

**Charité:** Sebastian Köhler^82^ SHU: Alison Metcalfe^48, 83^

**APHP:** Alain Verloes^84, 85^, Séverine Drunat^84, 85^, Delphine Heron^86, 87^, Cyril Mignot^86, 88^, Boris Keren^86^, Jean-Madeleine de Sainte Agathe^86^

**CHU Bordeaux:** Caroline Rooryck^89^, Didier Lacombe^89^, Aurelien Trimouille^90^

**Spain UDP:** Manuel Posada De la Paz^91^, Eva Bermejo Sánchez^91^, Estrella López Martín^91^, Beatriz Martínez Delgado^91^, F. Javier Alonso García de la Rosa^91^

**Ospedale Pediatrico Bambino Gesù, Rome:** Andrea Ciolfi^92^, Bruno Dallapiccola^92^, Simone Pizzi^92^, Francesca Clementina Radio^92^, Marco Tartaglia^92^

**University of Siena:** Alessandra Renieri^93–95^, Simone Furini^93, 94^, Chiara Fallerini^93, 94^, Elisa Benetti^93, 94^ Semmelweis University Budapest: Peter Balicza^96^, Maria Judit Molnar^96^ University of Ljubljana, Ales Maver^97^, Borut Peterlin^97^

**University of Lübeck:** Alexander Münchau^98^, Katja Lohmann^99^, Rebecca Herzog^98, 100^, Martje Pauly^98, 99^

**Val d’Hebron Barcelona:** Alfons Macaya^101, 102^, Ana Cazurro-Gutiérrez^101^, Belén Pérez-Dueñas^101^, Francina Munell^101^, Clara Franco Jarava^103, 104^, Laura Batlle Masó^105, 106^, Anna Marcé-Grau^101^, Roger Colobran^103, 104, 107^

**Hospital Sant Joan de Déu Barcelona:** Andrés Nascimento Osorio^108^, Daniel Natera de Benito^108^

**University of Freiburg:** Hanns Lochmüller^109–111^, Rachel Thompson^111^, Kiran Polavarapu^111^, Bodo Grimbacher^112–116^

**University of Oxford:** David Beeson^117^, Judith Cossins^117^

**Folkhälsan Research Centre:** Peter Hackman^118^, Mridul Johari^118^, Marco Savarese^118^, Bjarne Udd^118–120^

**University of Cambridge:** Rita Horvath^121^, Patrick F. Chinnery^121, 122^, Thiloka Ratnaike^123^, Fei Gao^121^, Katherine Schon^121, 124^

**Catalan Institute of Oncology, Barcelona:** Gabriel Capella^125^, Laura Valle^125^ KU Munich: Elke Holinski-Feder^126^, Andreas Laner^127^, Verena Steinke-Lange^126^ TU Dresden: Evelin Schröck^128^, Andreas Rump^128, 129^ Koç University: Ayşe Nazlı Başak^130^

**Ghent University Hospital:** Dimitri Hemelsoet^131, 132^, Bart Dermaut^132–134^, Nika Schuermans^132–134^, Bruce Poppe^132–134^, Hannah Verdin^133^

**University Hospital Meyer, Florence:** Davide Mei^135^, Annalisa Vetro^135^, Simona Balestrini^135, 136^, Renzo Guerrini^135^

**KU Leuven:** Kristl Claeys^137, 138^

**LUMC:** Gijs W.E. Santen^139^, Emilia K. Bijlsma^139^, Mariette J.V. Hoffer^139^, Claudia A.L. Ruivenkamp^139^

**Ludwig Boltzmann Institute for Rare and Undiagnosed Diseases, Vienna**: Kaan Boztug^140–144^, Matthias _Haimel_140-142

**Institute of Pathology and Genetics, Gosselies, Belgium:** Isabelle Maystadt^145, 146^

**Technical University Munich:** Isabell Cordts^147^, Marcus Deschauer^147^

**Neurology/Neurogenetics Laboratory University of Crete, Heraklion, Crete, Greece:** Ioannis Zaganas^148^, Evgenia Kokosali^148^, Mathioudakis Lambros^148^, Athanasios Evangeliou^149^, Martha Spilioti^150^, Elisabeth Kapaki^151^, Mara Bourbouli^151^

**IRCCS G. Gaslini:** Pasquale Striano^152, 153^, Federico Zara^153, 154^, Antonella Riva^153, 154^, Michele Iacomino^154, 155^, Paolo Uva^155^, Marcello Scala^152, 153^, Paolo Scudieri^153, 154^

**Cliniques universitaires Saint-Luc (CUSL):** Maria-Roberta Cilio^156^, Evelina Carpancea^156^, Chantal Depondt^157^, Damien Lederer^158^, Yves Sznajer^159^, Sarah Duerinckx^160^, Sandrine Mary^158^

**Institute of Human Genetics, University Hospital Essen:** Christel Depienne^161, 162^, Andreas Roos^111, 163, 164^

**University of Luxembourg:** Patrick May^165^

### Affiliations

1. Institute of Medical Genetics and Applied Genomics, University of Tübingen, Tübingen, Germany.
2. Centre for Rare Diseases, University of Tübingen, Tübingen, Germany.
3. NGS Competence Center Tübingen (NCCT), University of Tübingen, Tübingen, Germany.
4. Department of Neurodegeneration, Hertie Institute for Clinical Brain Research (HIH), University of Tübingen, Tübingen, Germany.
5. German Center for Neurodegenerative Diseases (DZNE), Tübingen, Germany.
6. Department of Neurology and Epileptology, Hertie Institute for Clinical Brain Research (HIH), University of Tübingen, Tübingen, Germany.
7. Department of Human Genetics, Radboud University Medical Center, Nijmegen, The Netherlands.
8. Donders Institute for Brain, Cognition and Behaviour, Radboud University Medical Center, Nijmegen, The Netherlands.
9. Department of Clinical Genetics, Maastricht University Medical Centre, Maastricht, the Netherlands.
10. Radboud Institute for Molecular Life Sciences, Nijmegen, The Netherlands.
11. Department of Internal Medicine and Radboud Center for Infectious Diseases (RCI), Radboud University Medical Center, Nijmegen, the Netherlands.
12. Center for Molecular and Biomolecular Informatics, Radboud University Medical Center, Nijmegen, the Netherlands.
13. Department of Neurology, Radboud University Medical Center, Nijmegen, The Netherlands.
14. Department of Genetics and Genome Biology, University of Leicester, Leicester, UK.
15. John Walton Muscular Dystrophy Research Centre, Translational and Clinical Research Institute, Newcastle University and Newcastle Hospitals NHS Foundation Trust, Newcastle upon Tyne, UK.
16. Primary Immunodeficiency Group, Translational and Clinical Research Institute, Newcastle University and Newcastle upon Tyne Hospitals NHS Foundation Trust, Newcastle upon Tyne, UK.
17. Division of Evolution, Infection and Genomics, School of Biological Sciences, Faculty of Biology, Medicine and Health, University of Manchester, Manchester M13 9WL, UK.
18. Manchester Centre for Genomic Medicine, St Mary’s Hospital, Manchester University Hospitals NHS Foundation Trust, Health Innovation Manchester, Manchester M13 9WL, UK.
19. Dijon University Hospital, Genetics Department, Dijon, France.
20. Dijon University Hospital, Centre of Reference for Rare Diseases: Development disorders and malformation syndromes, Dijon, France.
21. Inserm - University of Burgundy-Franche Comté, UMR1231 GAD, Dijon, France.
22. Dijon University Hospital, FHU-TRANSLAD, Dijon, France.
23. Dijon University Hospital, GIMI institute, Dijon, France.
24. University of Burgundy-Franche Comté, Dijon Economics Laboratory, Dijon, France.
25. University of Burgundy-Franche Comté, FHU-TRANSLAD, Dijon, France.
26. CNAG-CRG, Centre for Genomic Regulation (CRG), The Barcelona Institute of Science and Technology, Baldiri Reixac 4, Barcelona 08028, Spain.
27. Universitat Pompeu Fabra (UPF), Barcelona, Spain.
28. EURORDIS-Rare Diseases Europe, Sant Antoni Maria Claret 167 - 08025 Barcelona, Spain.
29. EURORDIS-Rare Diseases Europe, Plateforme Maladies Rares, 75014 Paris, France.
30. INSERM, US14 - Orphanet, Plateforme Maladies Rares, 75014 Paris, France.
31. Institut National de la Santé et de la Recherche Medicale (INSERM) U1127, Paris, France.
32. Centre National de la Recherche Scientifique, Unité Mixte de Recherche (UMR) 7225, Paris, France.
33. Unité Mixte de Recherche en Santé 1127, Université Pierre et Marie Curie (Paris 06), Sorbonne Universités, Paris, France.
34. Institut du Cerveau - ICM, Paris, France.
35. Ecole Pratique des Hautes Etudes, Paris Sciences et Lettres Research University, Paris, France.
36. Centre de Référence de Neurogénétique, Hôpital de la Pitié-Salpêtrière, Assistance Publique-Hôpitaux de Paris (AP-HP), Paris, France.
37. Hôpital de la Pitié-Salpêtrière, Assistance Publique-Hôpitaux de Paris (AP-HP), Paris, France.
38. Sorbonne Université, Inserm, Institut de Myologie, Centre de Recherche en Myologie, F-75013 Paris, France.
39. AP-HP, Centre de Référence de Pathologie Neuromusculaire Nord, Est, Ile-de-France, Institut de Myologie, G.H. Pitié-Salpêtrière, F-75013 Paris, France.
40. Institut de Myologie, Equipe Bases de données, G.H. Pitié-Salpêtrière, F-75013 Paris, France.
41. AP-HP, Unité Fonctionnelle de Cardiogénétique et Myogénétique Moléculaire et Cellulaire, G.H. Pitié-Salpêtrière, F-75013 Paris, France.
42. Department of Biology and Medical Genetics, Charles University Prague-2nd Faculty of Medicine and University Hospital Motol, Prague, Czech Republic.
43. Department of Paediatrics and Inherited Metabolic Disorders, First Faculty of Medicine, Charles University and General University Hospital in Prague, Prague, Czech Republic.
44. Department of Ophthalmology, First Faculty of Medicine, Charles University and General University Hospital in Prague, Prague, Czech Republic.
45. Centre for Paediatric Rheumatology and Autoinflammatory Diseases, Department of Paediatrics and Inherited Metabolic Disorders, 1st Faculty of Medicine, Charles University and General University Hospital in Prague, Czech Republic.
46. European Bioinformatics Institute, European Molecular Biology Laboratory, Wellcome Genome Campus, Hinxton, Cambridge, United Kingdom.
47. Jackson Laboratory for Genomic Medicine, Farmington, CT 06032, USA.
48. Florence Nightingale Faculty of Nursing, Midwifery & Palliative Care, King’s College, London, UK.
49. Society and Ethics Research, Connecting Science, Wellcome Genome Campus, Hinxton, UK.
50. Genomics England, Queen Mary University of London, Dawson Hall, EC1M 6BQ, London, UK.
51. MRC Centre for Neuromuscular Diseases and National Hospital for Neurology and Neurosurgery, UCL Queen Square Institute of Neurology, London, UK.
52. Department of Neuromuscular Diseases, UCL Queen Square Institute of Neurology, London, UK.
53. Department of Clinical and Movement Neurosciences, UCL Queen Square Institute of Neurology, University College London, WC1N 3BG.
54. Department of Clinical and Experimental Epilepsy, UCL Queen Square Institute of Neurology, London, UK.
55. Dementia Research Centre, Department of Neurodegenerative Disease, UCL Queen Square Institute of Neurology, London, UK.
56. Dubowitz Neuromuscular Centre, UCL Great Ormond Street Hospital, London, UK.
57. NIHR Great Ormond Street Hospital Biomedical Research Centre, London, United Kingdom.
58. Peripheral Neuropathy Research Group, University of Antwerp, Antwerp, Belgium.
59. Laboratory of Neuromuscular Pathology, Institute Born-Bunge, University of Antwerp, Antwerpen, Belgium.
60. Translational Neurosciences, Faculty of Medicine and Health Sciences, University of Antwerp, Belgium.
61. Neuromuscular Reference Centre, Department of Neurology, Antwerp University Hospital, Antwerpen, Belgium.
62. VIB-CMN, Applied and Translational Neurogenomics Group.
63. Dipartimento di Medicina di Precisione, Università degli Studi della Campania "Luigi Vanvitelli", Napoli, Italy.
64. Telethon Institute of Genetics and Medicine, Pozzuoli, Italy.
65. Istituto di Scienze Applicate e Sistemi Intelligenti "E.Caianiello" - ISASI -CNR.
66. Unit of Medical Genetics, Department of Medical Sciences, University of Ferrara, Italy.
67. Institute of Human Genetics, Medical Faculty, University of Bonn, Bonn, Germany.
68. Center for Hereditary Tumor Syndromes, University Hospital Bonn, Bonn, Germany.
69. i3S - Instituto de Investigação e Inovação em Saúde, Universidade do Porto, Portugal.
70. IPATIMUP - Institute of Molecular Pathology and Immunology of the University of Porto, Portugal.
71. Faculty of Medicine, University of Porto, Portugal.
72. Doctoral Programme in Biomedicine, Faculty of Medicine, University of Porto, Portugal.
73. Doctoral Programme in BiotechHealth, School of Medicine and Biomedical Sciences, University of Porto, Portugal.
74. Doctoral Programme in Computer Science, Faculty of Sciences, University of Porto, Portugal.
75. CHUSJ, Centro Hospitalar e Universitário de São João, Porto, Portugal.
76. Departament of Genetics, Faculty of Medicine, University of Porto, Portugal.
77. Faculty of Sciences, University of Porto, Portugal.
78. Department of Genetics, Genomics Coordination Center, University Medical Center Groningen, University of Groningen, Groningen, The Netherlands.
79. Department of Genetics, University Medical Center Groningen, University of Groningen, Groningen, The Netherlands.
80. ERN-GENTURIS.
81. ERN-RITA: European Reference Network for Immunodeficiency, Autoinflammatory, Autimmune and Paediatric Rheumatic diseases, Utrecht, Netherlands.
82. Ada Health GmbH, Karl-Liebknecht-Str. 1, 10178 Berlin, Germany.
83. College of Health, Well-being and Life-Sciences, Sheffield Hallam University, Sheffield, UK.
84. Dept of Genetics, Assistance Publique-Hôpitaux de Paris - Université de Paris, Robert DEBRE University Hospital, 48 bd SERURIER, Paris, France.
85. INSERM UMR 1141 "NeuroDiderot", Hôpital Robert DEBRE, Paris, France.
86. Department of Genetics, Assistance Publique-Hôpitaux de Paris - Sorbonne Université, Pitié-Salpêtrière University Hospital, 83 Boulevard de l’Hôpital, Paris, France.
87. Reference center of rare diseases "intellectuel disability of rare causes", Paris, France.
88. Institut du Cerveau (ICM), UMR S 1127, Inserm U1127, CNRS UMR 7225, Sorbonne Université, 75013, Paris, France.
89. Univ. Bordeaux, MRGM INSERM U1211, CHU de Bordeaux, Service de Génétique Médicale, F-33000 Bordeaux, France.
90. Laboratoire de Génétique Moléculaire, Service de Génétique Médicale, CHU Bordeaux – Hôpital Pellegrin, Place Amélie Raba Léon, 33076 Bordeaux Cedex, France.
91. Institute of Rare Diseases Research, Spanish Undiagnosed Rare Diseases Cases Program (SpainUDP) & Undiagnosed Diseases Network International (UDNI), Instituto de Salud Carlos III, Madrid, Spain.
92. Molecular Genetics and Functional Genomics, Ospedale Pediatrico Bambino Gesù, IRCCS, Rome, Italy.
93. Med Biotech Hub and Competence Center, Department of Medical Biotechnologies, University of Siena, Italy.
94. Medical Genetics, University of Siena, Italy.
95. Genetica Medica, Azienda Ospedaliero-Universitaria Senese, Italy.
96. Institute of Genomic Medicine and Rare Diseases, Semmelweis University, Budapest, Hungary.
97. Clinical Institute of Genomic Medicine, University Medical Centre Ljubljana, Slovenia.
98. Institute of Systems Motor Science, University of Lübeck, Ratzeburger Allee 160, 23562, Lübeck, Germany.
99. Institute of Neurogenetics, University of Lübeck, Ratzeburger Allee 160, 23562, Lübeck, Germany.
100. Department of Neurology, University Hospital Schleswig Holstein, Ratzeburger Allee 160, 23562, Lübeck, Germany.
101. Pediatric Neurology Research Group, Vall d’Hebron Research Institute, Universitat Autònoma de Barcelona, Barcelona, Spain.
102. Institute of Neuroscience, Universitat Autònoma de Barcelona, Barcelona, Spain.
103. Diagnostic Immunology Research Group, Vall d’Hebron Research Institute (VHIR), Barcelona, Spain.
104. Immunology Division, Genetics Department. Vall d’Hebron University Hospital (HUVH), Barcelona, Spain.
105. Infection in Immunocompromised Pediatric Patients Research Group, Vall d’Hebron Research Institute (VHIR), Barcelona, Spain.
106. Pediatric Infectious Diseases and Immunodeficiencies Unit, Vall d’Hebron University Hospital (HUVH),Barcelona, Spain.
107. Immunology Unit. Department of Cell Biology, Physiology and Immunology. Autonomous University of Barcelona (UAB), Bellaterra, Spain.
108. Neuromuscular Disorders Unit, Department of Pediatric Neurology. Hospital Sant Joan de Déu, Barcelona, Spain
109. Department of Neuropediatrics and Muscle Disorders, Medical Center, Faculty of Medicine, University of Freiburg, Freiburg, Germany.
110. Centro Nacional de Análisis Genómico (CNAG-CRG), Center for Genomic Regulation, Barcelona Institute of Science and Technology (BIST), Barcelona, Spain.
111. Children’s Hospital of Eastern Ontario Research Institute, University of Ottawa, Ottawa, Canada.
112. Institute for Immunodeficiency, Center for Chronic Immunodeficiency (CCI), Medical Center, Faculty of Medicine, Albert-Ludwigs-University of Freiburg, Germany.
113. Clinic of Rheumatology and Clinical Immunology, Center for Chronic Immunodeficiency (CCI), Medical Center, Faculty of Medicine, Albert-Ludwigs-University of Freiburg, Germany.
114. DZIF – German Center for Infection Research, Satellite Center Freiburg, Germany.
115. CIBSS – Centre for Integrative Biological Signalling Studies, Albert-Ludwigs University, Freiburg, Germany.
116. RESIST – Cluster of Excellence 2155 to Hanover Medical School, Satellite Center Freiburg, Germany.
117. Nuffield Department of Clinical Neurosciences, University of Oxford, UK.
118. Folkhälsan Research Centre and Medicum, University of Helsinki, Helsinki, Finland.
119. Tampere Neuromuscular Center, Tampere, Finland.
120. Vasa Central Hospital, Vaasa, Finland.
121. Department of Clinical Neurosciences, University of Cambridge, Cambridge, UK.
122. Medical Research Council Mitochondrial Biology Unit, University of Cambridge, Cambridge, UK.
123. Department of Paediatrics, University of Cambridge, Cambridge, UK.
124. East Anglian Medical Genetics Service, Cambridge University Hospitals NHS Foundation Trust, Cambridge, UK.
125. Bellvitge Biomedical Research Institute (IDIBELL), Barcelona, Spain.
126. Medizinische Klinik und Poliklinik IV – Campus Innenstadt, Klinikum der Universität München, Munich, Germany.
127. MGZ - Medical Genetics Center, Munich, Germany.
128. Institute of Clinical Genetics, University Hospital Carl Gustav Carus, Technical University Dresden, Dresden, Germany.
129. Center for Personalized Oncology, University Hospital Carl Gustav Carus, Technical University Dresden, Dresden, Germany.
130. Koç Universıty,School of Medicine, Translational Medicine Research Center, KUTTAM-NDAL Istanbul Turkey.
131. Dpt. of Neurology, Ghent University Hospital, Ghent, Belgium.
132. Program for Undiagnosed Rare Diseases (UD-PrOZA), Ghent University Hospital, Ghent, Belgium.
133. Center for Medical Genetics, Ghent University Hospital, Ghent, Belgium.
134. Department of Biomolecular Medicine, Faculty of Medicine and Health Sciences, Ghent University, Ghent, Belgium.
135. Neuroscience Department, Children’s Hospital A. Meyer-University of Florence, 50139, Florence, Italy.
136. Department of Clinical and Experimental Epilepsy, UCL Queen Square Institute of Neurology, and Chalfont Centre for Epilepsy, Gerrard Cross, UK.
137. Department of Neurology, University Hospitals Leuven, Leuven, Belgium.
138. Laboratory for Muscle Diseases and Neuropathies, Department of Neurosciences, and Leuven Brain Institute (LBI), KU Leuven - University of Leuven, Leuven, Belgium.
139. Department of Clinical Genetics, Leiden University Medical Center, Leiden, The Netherlands.
140. Ludwig Boltzmann Institute for Rare and Undiagnosed Diseases, Vienna, Austria.
141. St. Anna Children’s Cancer Research Institute (CCRI), Vienna, Austria.
142. CeMM Research Center for Molecular Medicine of the Austrian Academy of Sciences, Vienna, Austria.
143. Department of Pediatrics and Adolescent Medicine, Medical University of Vienna, Vienna, Austria.
144. 144. St. Anna Children’s Hospital, Department of Pediatrics and Adolescent Medicine, Medical University of Vienna, Vienna, Austria.
145. Centre de Génétique Humaine, Institut de Pathologie et de Génétique, Gosselies, Belgium.
146. Département de Médecine, Université de namur (Unamur), Namur, Belgique.
147. Department of Neurology, Klinikum rechts der Isar, Technical University Munich, Munich, Germany.
148. Neurology / Neurogenetics Laboratory University of Crete, Heraklion, Crete, Greece.
149. Aristotle University of Thessaloniki, Thessaloniki, Greece.
150. 1st Department of Neurology, Aristotle University of Thessaloniki, University General Hospital of Thessaloniki, AHEPA, Thessaloniki, Greece.
151. Neurochemistry and Biomarker Unit, 1st Department of Neurology, School of Medicine, National and Kapodistrian University of Athens, Eginition Hospital, Athens, Greece.
152. Pediatric Neurology and Muscular Disease Unit, IRCCS Istituto Giannina Gaslini, Genoa, Italy.
153. Department of Neurosciences, Rehabilitation, Ophthalmology, Genetics, Maternal and Child Health, University of Genoa, Genoa, Italy.
154. Unit of Medical Genetics, IRCCS Istituto Giannina Gaslini, Genoa, Italy.
155. Clinical Bioinformatics, IRCCS Istituto Giannina Gaslini, Genoa, Italy.
156. Pediatric Neurology Department, Saint-Luc University Hospital, Université Catholique de Louvain, Brussels, Belgium.
157. Neurology Department, Erasme Hospital, Université Libre de Bruxelles, Bruxelles, Belgium.
158. Institute of Pathology and Genetics, Charleroi, Belgium.
159. Human Genetics Department, Saint-Luc University Hospital, Université Catholique de Louvain, Brussels, Belgium.
160. Institute of Interdisciplinary Research in Human and Molecular Biology, Human Genetics, IRIBHM, Université Libre de Bruxelles, Brussels, Belgium.
161. Institute of Human Genetics, University Hospital Essen, University Duisburg-Essen, Essen, Germany.
162. Institut du Cerveau et de la Moelle épinière (ICM), Sorbonne Université, UMR S 1127, Inserm U1127, CNRS UMR 7225, F-75013 Paris, France.
163. Department of Pediatric Neurology, Developmental Neurology and Social Pediatrics, Children’s Hospital University of Essen, Essen, Germany.
164. Department of Neurology, Heimer Institute for Muscle Research, University Hospital Bergmannsheil, Ruhr-University Bochum, 44789 Bochum, Germany.
165. Luxembourg Centre for Systems Biomedicine, University of Luxembourg, Esch-sur-Alzette, Luxembourg.

