## Supplementary Materials for "Comprehensive reanalysis for CNVs in ES data from unsolved rare disease cases results in new diagnoses"


Title ... 2

Authors ... 2

Author Contributions ... 2

Supplementary Methods ... 3

CNV Detection ... 3

Alignment and definition of capture regions of interest ... 3

ClinCNV Workflow ... 3

Conifer Workflow ... 5

ExomeDepth Workflow ... 6

CNV Classes ... 7

CNV Visualisation ... 8

Diagnostic interpretation ... 8

ERN EURO-NMD ... 9

ERN GENTURIS ... 10

ERN ITHACA ... 10

ERN RND ... 11

References ... 12

Supplementary Figures ... 13

Supplementary Figure 1 ... 13

Supplementary Figure 2 ... 14

Supplementary Figure 3 ... 15

Supplementary Figure 4 ... 16

Supplementary Tables (see Excel file) ... 16

### Title

*Comprehensive reanalysis for CNVs in ES data from unsolved rare disease cases results in new diagnoses*

### Authors

German Demidov[1,2]*‡, Burcu Yaldiz[3,4]*, José Garcia-Pelaez[5,6,7]*, Elke de Boer[3,8,9]*, Nika Schuermans[10]*, Liedewei Van de Vondel[11,12]*, Ida Paramonov[13], Lennart F. Johansson[14], Francesco Musacchia[15,16], Elisa Benetti[17], Gemma Bullich[13], Karolis Sablauskas[3,18], Sergi Beltran[13,19,20], Christian Gilissen[3], Alexander Hoischen[3,21,22], Stephan Ossowski[1,2], Richarda de Voer[3,23], Katja Lohmann[24], Carla Oliveira[5,6,7], Ana Topf[25], Lisenka E.L.M. Vissers[3,8], the Solve-RD Consortia, Steven Laurie[13]*‡

### Author Contributions

* These authors contributed equally to this work

‡ Corresponding authors

The authors declare no conflicts of interest.

### Supplementary Methods

#### CNV Detection

##### Alignment and definition of capture regions of interest

Sequencing data was submitted in BAM, CRAM, or FastQ format. Where data was submitted in BAM or CRAM format it was reconverted to FastQs at read-group level prior to being realigned to the hs37d5 human genome reference version, as used in phase 2 of the 1000 genomes project[1] with BWA-MEM[2] (v0.7.8-r455). As GC-rich enrichment targets are known to amplify poorly, resulting in unreliable CNV calling[3], the GC-content for each target in each enrichment kit was calculated and any targets in which the GC-content was >80% were removed from the corresponding target BED file prior to CNV calling. This resulted in the removal of <0.5% of target regions per kit. Ensembl version 75 was used for gene and transcript definition.

##### ClinCNV Workflow

Analysis was performed separately for experiments generated by different exome enrichment kits. Initially, ClinCNV calculates the average read coverage of targeted regions of the enrichment kit divided into 120bp windows. As the first step of preprocessing, coverage is corrected for GC-content and library size for each sample individually. Following normalisation, systematically poorly covered regions (i.e. where 90% of samples had a normalised coverage <0.3) were excluded, followed by the application of variance stabilisation of read counts (square root transformation). To ameliorate the potential impact of batch effects on coverage calculation, samples were further clustered based on their global coverage profiles. In generating these clusters, target regions in the top and bottom quintiles for variance were excluded to minimise the potential impact of polymorphic regions on cluster generation, and coverage profiles smoothed using the rolling median. Uniform manifold approximation and projection (UMAP)[4] was performed for the mapping of smoothed coverage profiles. Samples were clustered into subgroups with a minimum size of 15 using dbscan[5]. Finally, the coverage of each 120bp window was normalised using the median of coverages within the cluster. Different potential copy numbers are modelled using the theoretical expected value and estimated variance, and the log-likelihood of normalised coverage under different expected copy-number models is calculated for each window. Calling is performed analogously to Circular Binary Segmentation[6] using a Maximum Subarray Sum algorithm[7] *i.e.* the segment with the highest evidence supporting an alternative copy-number to that of the model is identified at each step of the segmentation, rather than the segment with the largest difference in mean.

Resulting CNV calls were filtered according to measures of within kit allele frequency of the CNV and the noisiness of the coverage at the CNV site, requiring a minimum log-likelihood ratio of 20 to be considered worthy of biological interpretation. A robust regression model is fitted, taking the 75% percentile rank of the per-chromosome number of CNVs as a response variable, and median read depth, enrichment kit, and predicted ancestry determined using SampleAncestry (https://github.com/imgag/ngs-bits/blob/master/doc/tools/SampleAncestry/index.md) as predictors. A sample was assessed as QC failed if the response variable was outwith the 99.5% prediction interval of the regression. The 75% percentile of the per-chromosome number of CNVs was chosen to overcome cases where long CNVs may have been segmented into many separate calls and thus an otherwise good sample could be falsely identified as QC failed if only the total number of CNV calls was used as a response. Where parents of a case were available (*i.e*. family trios), copy-number information from the parents was also provided to assist in interpretation, and to confirm if CNVs represented *de novo* events.

#### Conifer Workflow

Conifer[8] (http://conifer.sourceforge.net/) uses Singular Value Decomposition (SVD) to identify rare CNVs from exome sequencing data. Samples with similar read lengths were analysed in the same batch, and sex-specific sample pools were created for generating accurate X-Chromosome calls. RPKM (Reads Per Kilobase per Million mapped reads) values were calculated independently by enrichment kit for all corresponding targets. Following SVD to identify biases in coverage introduced by batch effects, 3 to 15 components were removed from each group based on manual inspection of the inflection points of scree plots generated by the program.

Within each analysis batch, if all experiments had less than 30 calls, the results were considered ready for further filtering. On the contrary where any experiment in a batch had more than 30 calls, then if the median number of calls per experiment in the batch was less than 10, any experiment with more than 30 calls was discarded as failing QC, and the results from the remaining experiments were considered ready for filtering. However, if the median number of calls within the batch was more than 10 per experiment then the SVD value was increased, and the batch analysis rerun, until either all experiments had less than 30 calls, or the median number of calls was less than 10, at which point any experiment with more than 30 calls was discarded as described above. CNVs with an SVD-ZRPKM value greater than 1.75 or less than -1.75 were considered as *bona fide* duplication or deletion calls, respectively, worthy of biological interpretation. Conifer does not provide any guidance as to the exact copy number identified at a particular locus and provides no further indicators of the quality of a detected event other than the SVD-RPKM metric.

#### ExomeDepth Workflow

ExomeDepth[9], applies a beta-binomial model to the genome-wide distribution of read depth data, aiming to compare a test sample to a similar reference set selected by the tool. For the implementation of the ExomeDepth workflow, the generation of read count data was separated from that of identifying candidate CNVs. Thus for each experiment, read depth was initially calculated for all targets of the respective capture kit, and stored as a Bioconductor iRanges object[10]. In the second step, all iRanges objects from experiments generated using the same enrichment kit were analysed as a batch to generate raw CNV call sets. In this second step ExomeDepth automatically identifies an independent background reference set for each test sample by selecting the most closely correlated samples in terms of coverage from within the batch. Copy-Number prediction is provided by the ratio of observed/expected reads over a set of targets. We interpreted these ratios in diploid chromosomes as follows:

- O/E ratio <0.10 - Likely Homozygous Deletion i.e. Copy Number (CN) = 0
- 0.10< O/E ratio <0.75 - Likely Heterozygous Deletion; CN=1
- 0.75< O/E ratio <1.25 - Likely Copy Number neutral; CN=2 *i.e.* No CNV to report
- 1.25< O/E ratio <1.75 - Likely Heterozygous Duplication; CN=3
- 1.75< O/E ratio <2.25 - CN=4
- O/E ratio >2.25 - CN OTHER

ExomeDepth provides two indicators of quality. The first is a sample-level indicator of the correlation between the test sample and the background reference, which should be >0.97 for the results to be regarded as reliable. Secondly, regarding call quality, ExomeDepth provides a Bayes Factor (BF) based on the ratio of observed/expected reads over a set of apparently copy-number variant targets. Experiments with a correlation <0.97 were considered as failing QC, and any calls with a BF<0.15 were discarded as being unreliable.

#### CNV Classes

To aid downstream interpretation, each CNV call was categorised into one of six classes.

1) Putative CNVs longer than 500kb in length were initially identified regardless of the presence of absence of genes of interest in the ERN gene lists. The recent release of large CNVs catalogues such as DECIPHER , as well as the presence of a large number of case reports with chromosomal changes of this size and larger, allowed us to hypothesise that such variants could be interpreted successfully, even if the reported phenotypes of the patients exhibiting such variants may differ from the phenotypes expected for affected genes.
2) Homozygous deletions are generally rare, and the presence of a homozygous deletion needs to be interpreted very cautiously due to potentially incorrect enrichment kit reporting, or poor-quality library preparation. An important indicator that a putative homozygous deletion call is likely to be *bona fide* is the consanguinity status of the patient.
3) Heterozygous CNVs occurring in genes with a described autosomal-dominant mode of inheritance reported in OMIM.
4) Duplications with apparent copy number >3. These may represent cases where alleles on both chromosomes are duplicated, or cases where only the allele on one chromosome has been duplicated but multiple times.
5) Gonosomal CNVs. As gonosomal CNVs require a mixed workflow depending on the sex of the participant, a separate set of calls was generated for CNV calls on chromosomes X and Y. In the case of the Y-Chromosome, only “Long” CNVs that would fall into category 1 above were reported for interpretation, since there were no genes of interest on the Y-Chromosome on any of the ERN gene lists.
6) Potential compound heterozygote SNV/CNV “double-hits”. For a short list of experiments in which a single candidate SNV had been identified by the Solve-RD

SNV working group, which was either listed in ClinVar as Pathogenic/Likely Pathogenic or predicted to have a high impact in a gene of interest, affecting an individual where the mode of inheritance was suspected to be recessive, (see Laurie et al, 2023, under review) we investigated whether a potentially pathogenic CNV affecting the second allele of the same gene could explain the case as a compound heterozygote.

#### CNV Visualisation

To provide support for interpretation of the technical validity of CNV calls, screenshots for regions containing CNV calls were generated automatically using the Integrative Genomics Viewer[11] (IGV), incorporating a variety of custom-built tracks (see **Supplementary Figure 3**). These included call tracks for each of the three callers in SEG format, normalised coverage tracks for ClinCNV and Conifer, beta-allele frequency, BAM DoC, Institute of Medical Genetics and Applied Genomics (Tübingen) in-house polymorphic CNV regions, and gene tracks from RefSeq genes, ERN candidate genes, and DECIPHER microdeletion and duplication syndromes[12].

For each CNV returned for interpretation, we generated IGV screenshots of both the whole sample (chr1-22 and chrX/Y) to allow evaluation of overall sample quality, and the region around the individual CNV (+/-10kb). Specifically in the case of long CNVs, the observation of clear deviations from the expected ratio of 50/50 in beta-allele frequencies provided strong additional support of variant validity. For rare cases in which a signal of unusual read pairing was observed, suggesting that a breakpoint may have been captured, a screenshot was generated including the suspected breakpoint.

#### Diagnostic interpretation

To facilitate diagnostic interpretation, we used AnnotSV[13] (version 3.0.7) to add a range of useful annotations to the reports returned to Clinical experts for interpretation as listed in

**Supplementary Table 2**. Diagnostic interpretation was undertaken by expert clinicians and clinical researchers from the respective ERNs. Each ERN prioritised the calls for further investigation according to their own strategy, based on their expert knowledge of underlying disease mechanisms in their respective patients. Some annotations, such as that of the ENCODE blacklist for high-signal regions were used to quickly discard overlapping CNVs by all ERNs, whereas other information, such as evidence of consanguinity, provided further support that homozygous deletions were likely to be relevant in affected cases. For the interpretation of heterozygous deletions, pLI scores from GnomAD[14], and haploinsufficiency gene lists from the DDD project[15], aided interpretation. The full workflow is illustrated in **Supplementary Figure 4**.

#### ERN EURO-NMD

The filtering strategy undertaken by EURO-NMD was determined per analysis (see section 'CNV filtering'). In general, a balance had to be upheld whereby submitting clinical researchers would interpret as many CNVs as possible while maintaining a feasible interpretation load. Thus the following analyses were shared directly given the relative number of CNVs to be analysed: homozygous deletions, high copy number duplications, gonosomal CNVs, and potential compound heterozygote second hits, whereas heterozygous CNVs were split between CNVs of copy number one (CN1, i.e. deletions) and those of copy number three (CN3 i.e. duplications).

For CN1, CNVs for genes with DDD Haploinsuffiency scores > 90 or a GnomAD pLi < 0.1 were discarded, as these indicate that the gene is likely tolerant of heterozygous deletions. For both CN1 and CN3, CNVs identified through ClinCNV with a loglikelihood < 30 were discarded, as these are likely false positives. CNVs identified in genes only known to have recessive inheritance patterns were discarded, as were CNVs reported in Conrad *et al*[16]. For long CNVs, CNVs found in the Encode blacklist were discarded. Following these filtering steps, experts from the submitting groups applied a phenotype-first approach. If the phenotype could potentially match with the gene affected by the CNV call, IGV tracks were checked to evaluate the likelihood of the called CNV being a true CNV.

#### ERN GENTURIS

Due to the small size of the ERN GENTURIS cohort, and the short gene list, only limited further filtering of calls was necessary. No additional filters were applied to call sets from Conifer. In the case of heterozygote deletions and duplications, specific filtering criteria were applied separately for ClinCNV and ExomeDepth. For ClinCNV, we first interpreted all events identified by more than one tool, independent of the ClinCNV loglikelihood value. After this, we proceeded to analyse all events called only by ClinCNV with a loglikelihood of at least 20. For ExomeDepth, we first interpreted all events called by more than one tool, independently of the Bayes Factor (BF), and subsequently considered events called only by ExomeDepth with a BF of at least 15. For long CNVs, we first discarded all those events found in the encode blacklist and analysed the rest. For all datasets, following IGV visualization, only CNVs observed to be rare in control populations were considered for further interpretation.

#### ERN ITHACA

For ERN ITHACA, as a first step, we discarded variants that were annotated to have low QC, had been previously annotated as benign, or occurred in regions on the Encode Blacklist, as provided by the AnnotSV annotation. Additionally, to reduce the proportion of false positives, we discarded deletions shorter than 10kb and duplications shorter than 20kb in length, with the exception of homozygous deletion calls and variants in parent-offspring trios identified as being *de novo* by ClinCNV. Following this, visual inspection of each of the remaining CNV calls in IGV images was undertaken to assess technical validity, using reads and coverage supporting the call and B-allele frequency. Based on this visual assessment, apparently real biological CNVs were defined. For detailed clinical interpretation, prioritisation was subsequently guided by genes present on the ERN ITHACA gene list with a disease-association validity score ≥3, see Laurie et al, 2023 (under review), consistent with the expected mode of inheritance. Of note, CNVs ≥200 kb were also investigated regardless of the presence or absence of a gene on the ERN ITHACA gene list, given the prior knowledge of large CNVs being involved in ITHACA-associated phenotypes. All CNVs passing the above criteria were returned to the submitting groups from DITF-ITHACA, for diagnostic interpretation based on the clinical relevance to the phenotype observed in the affected individual.

#### ERN RND

The filtering strategy of ERN RND was predominantly based on tool-specific metrics. In general, the goal was to exclude calls with a high likelihood of being false positives. For ClinCNV we discarded all calls with a loglikelihood <30 and fist prioritised calls with a loglikelihood >200. As Conifer provides no metrics for filtering, all Conifer calls were analysed. For ExomeDepth, we discarded all calls affecting less than three targets and those with a Bayes factor <30, unless there was an overlapping CNV identified by one of the other tools. Following these filtering steps, the clinical researchers who submitted the case applied a phenotype-first approach. If the phenotype could potentially match that of the called CNV, IGV tracks were checked visually to evaluate the likelihood that the called CNV was *bona fide*.

### Supplementary Figures

#### Supplementary Figure 1

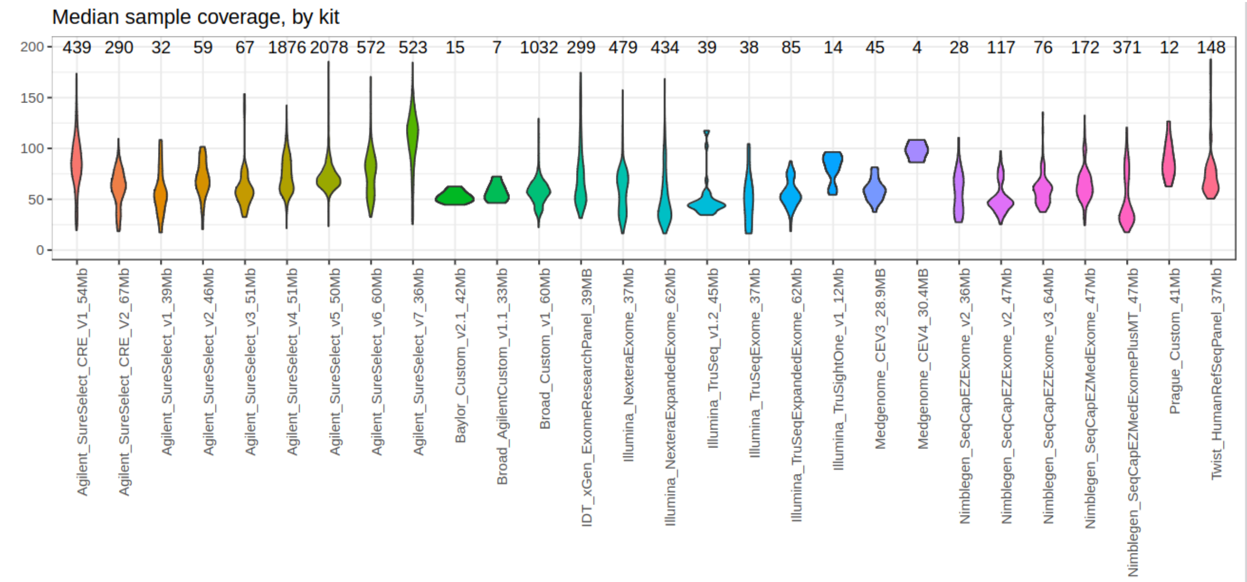


**Supplementary Figure 1.** Violin plot of median depth of coverage by kit for 9,351 ES experiments pertaining to 28 different enrichment kits. The number of experiments pertaining to each kit is shown above the plots. Coverage is shown on the Y-axis. Thickness of the plotted shape indicates the proportion of experiments which have a particular coverage.

#### Supplementary Figure 2

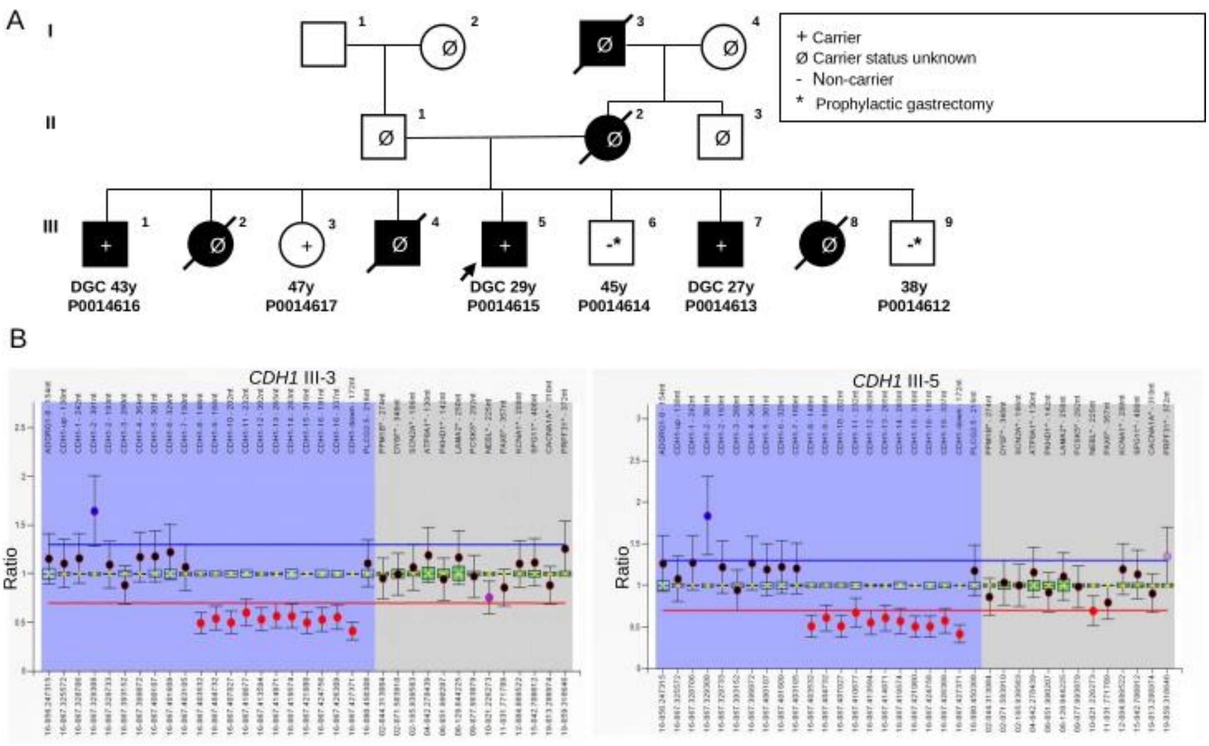


**Supplementary Figure 2.** Family pedigree and MLPA confirmation results for a Mexican family extensively affected by Hereditary Gastric Cancer. **A)** Family tree of the family of proband P0014615 (represented by an arrow). Exome Sequencing data from six individuals of the family was submitted to Solve-RD for re-analysis, following prior analysis in 2015 for both SNVs and CNVs which retrieved a negative result. Three of the sequenced family members were affected by diffuse gastric cancer (DGC, black symbols: P0014616, P0014615, P0014613) while the other three were unaffected (P0014617, P0014614, P0014612). Individual III-3 (P0014617) is currently a healthy carrier, perhaps due to the incomplete penetrance reported for *CHD1*. The age shown below affected individuals indicates age of disease onset, while that below healthy individuals represents their current age. **B)** MLPA validation results using SALSA MLPA-Probemix P083 *CDH1* (MRC Holland) in the healthy-carrier III-3, and in the proband, III-5. A ratio above the blue line indicates elevated number of copies, while a ratio below the red line indicates a decrease in copy number. The shaded blue area represents position of probes for *CDH1* and two neighbouring genes while the grey area represents reference probes.

#### Supplementary Figure 3

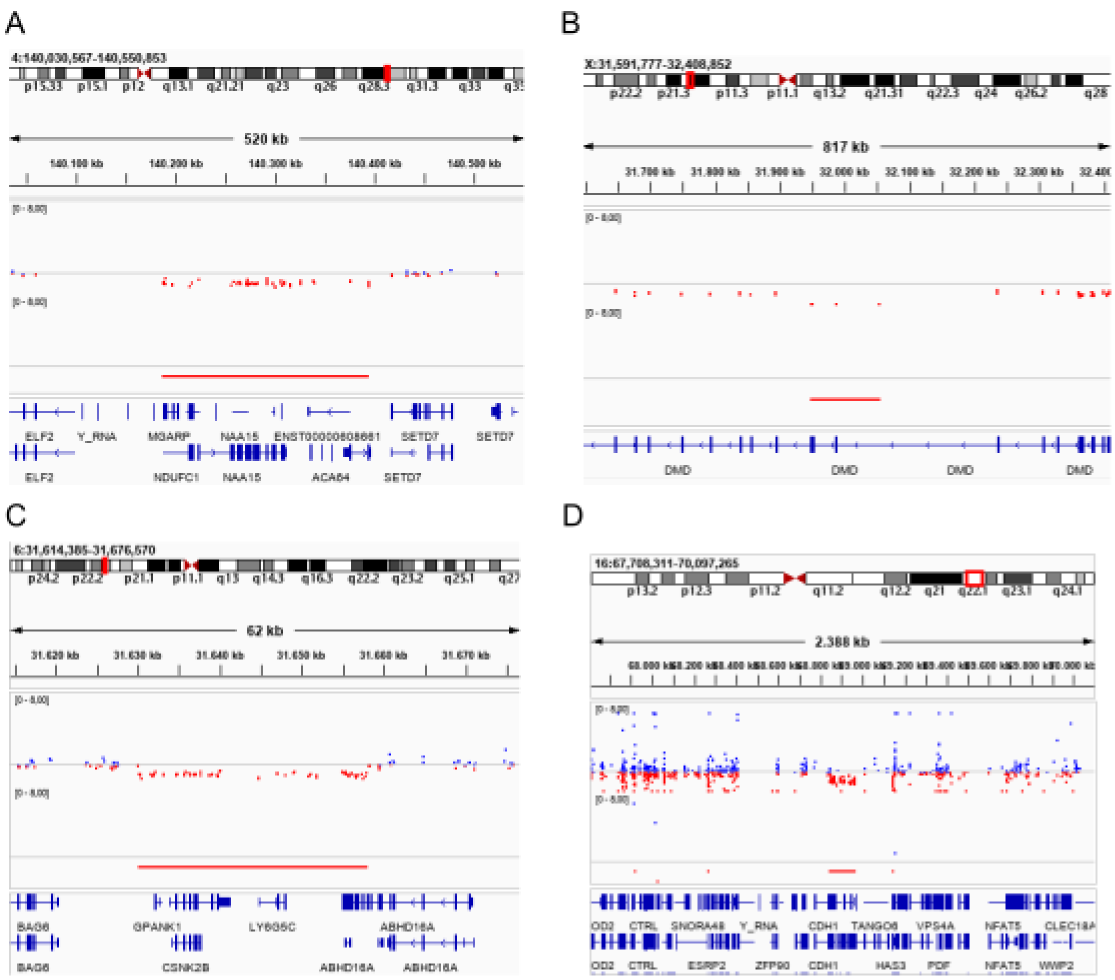


**Supplementary Figure 3.** IGV screenshots corresponding to the four illustrative newly diagnosed individuals described in the main text, one from each ERN. **A)** RND: Heterozygous deletion spanning *NAA15*, in an individual with intellectual disability, which was found to be inherited from her paucisymptomatic mother. **B)** EURO-NMD: Hemizygous deletion of exons 45-47 of *DMD* resulting in Becker Muscular Dystrophy. **C)** ITHACA: Heterozygous *de novo* deletion spanning *CSNK2B*, resulting in POBINDS **D)** GENTURIS: Inherited heterozygous deletion affecting *CDH1* and *TANGO6*, resulting in autosomal dominant HDGC. Images show customised coverage tracks and the position of the identified CNV (red bar). Blue dots above the midline indicate elevated coverage, while red dots below the line indicate reduced coverage. The position of genes is indicated at the bottom of the image, while the chromosomal position is indicated at the top of the image.

### Supplementary Figure 4

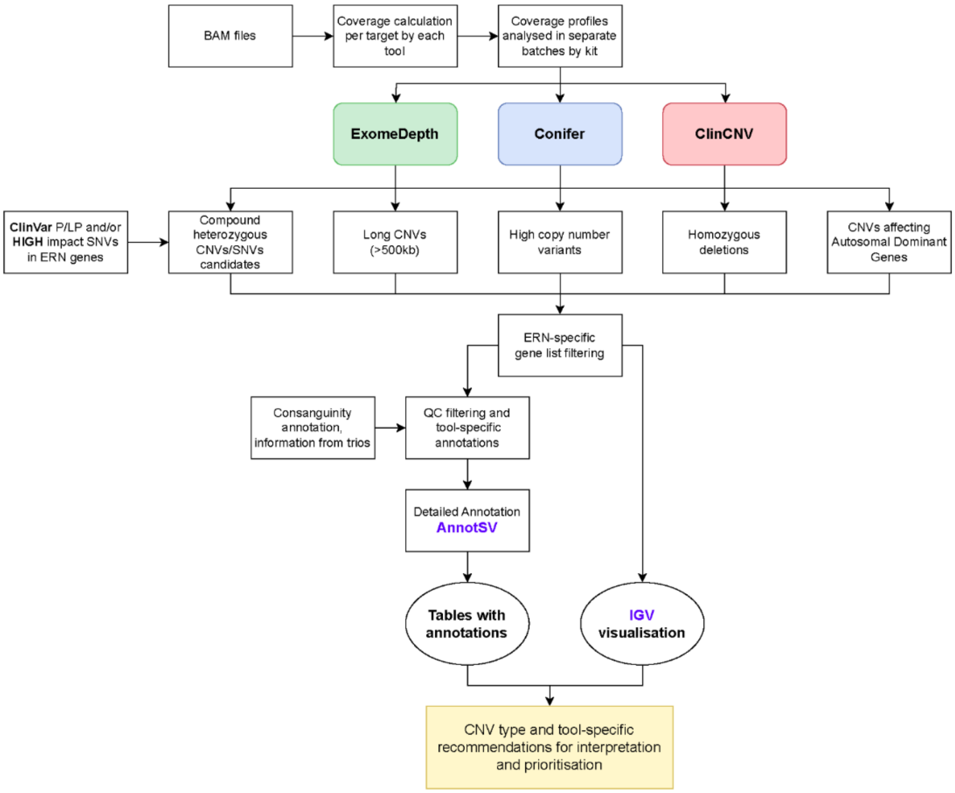


**Supplementary Figure 4**. Workflow used for CNV calling, filtering, and annotation, prior to returning to calls to clinical experts for interpretation.

### Supplementary Tables (see Excel file)

**Supplementary Table 1.** Curated ERN gene lists.

**Supplementary Table 2.** Annotations added to CNV calls to aid variant interpretation.

**Supplementary Table 3.** Number of families and affected individuals analysed per ERN, and number of families and affected individuals with at least one CNV requiring interpretation.

**Supplementary Table 4.** Summary statistics regarding 7,849 CNVs initially returned for interpretation.

**Supplementary Table 5.** Summary statistics regarding the length of 3,487 duplications and 4,362 deletions returned for interpretation.
